# Myocardial-derived small extracellular vesicles spontaneously released from living myocardial slices under biomimetic culture conditions regulate contractility and cardiac remodelling

**DOI:** 10.1101/2024.02.09.24302607

**Authors:** L. Nicastro, A. Lal, A. Kyriakou, S. Kholia, R. Nunez Toldra, B. Downing, F. Kermani, M. Anwar, F. Martino, D. Chokron, P. Sarathchandra, M. Sarkar, C. Emanueli, C.M. Terracciano

## Abstract

**BACKGROUND:** Small extracellular vesicles (sEVs) released in the cardiac microenvironment are reported to regulate cardiac remodelling, partially via microRNA transfer. Harvesting sEVs produced exclusively from the myocardium remains challenging and a solid research platform for sEV cardiovascular testing needs to be established. Organotypic living myocardial slices (LMS) allow to mimic cardiac disease and to record electrophysiological responses to biological and pharmacological stimuli. This study aims at understanding how cardiac sEVs obtained from donor and failing human LMS and rat LMS under physiological or heart failure-mimicking conditions impact myocardial function and remodelling.

**METHODS & RESULTS:** Human LMS were obtained from the left ventricle (LV) of human donor non-failing and end-stage failing hearts and cultured at 2.2 µm sarcomere length (SL). Rat LV LMS from healthy Sprague-Dawley rats were cultured at a preload of 2.2 or 2.4 µm SL, to recapitulate physiological load and overload, respectively. Following 48-hours biomimetic culture, sEVs were isolated from the culture media by size exclusion chromatography and characterized for their size, concentration, and expression of exosome markers. LMS from human failing hearts presented impaired contractility (P<0.05 vs donor-LMS), which was improved by application of donor heart-derived sEVs at 15 and 20% stretch. Whilst rat overloaded sEVs did not alter the force production of physiological LMS, physiological sEVs significantly increased the active force and decreased their passive force. In rat LMS, 1×10^8^ physiological EVs/slice restored the contractility of overloaded slices, reduced apoptosis, fibrosis-related gene expression and promoted angiogenesis. microRNAs analysis showed significant upregulation of miR-23a-3p and miR-378a-3p in rat physiological sEVs. Finally, to test whether sEVs have a direct effect on cardiomyocytes, we applied sEVs on cultured induced pluripotent stem cell-derived cardiomyocytes (iPSC-CMs). sEVs did not affect the contractility of iPSC-CM monoculture but increased the contractility of iPSC-CM co-cultured with human microvasculature endothelial cells (MVECs).

**CONCLUSIONS:** Cardiac sEVs isolated from healthy hearts increase the contractility of failing LMS. This effect is associated with, and possibly brought about by, a combination of inhibition of apoptosis, reduction of fibrosis and increased microvascular density, and could involve the transfer of sEV-microRNA into myocardial cells. Our data support the hypothesis that the sEV inotropic action is mediated by endothelial cells.

## Introduction

Increased mechanical load caused by either excessive pressure or volume in the left ventricle is an important driver of cardiac remodelling. Multiple changes take place during remodelling caused by mechanical overload, such as development of hypertrophy, myocardial cells death, fibrosis, and blood vessel rarefaction. Over time these changes lead to ventricular dysfunction and ultimately heart failure^1^.

During both physiology and disease, cell-cell communication is responsible for driving cellular and molecular changes to respond to changes in the environment ^2,3^. Numerous mechanisms of cell-cell communication, from direct contact to the release of soluble paracrine and endocrine factors, have been extensively investigated and are known to be important mediators of both cardiac homeostasis and remodelling.

Extracellular vesicles (EVs) encompass several types of nanosized and micro-sized vesicles with a lipid bilayer membrane and bioactive cargoes, including nucleic acids, proteins, lipids and metabolites ^4^. Small EVs (sEVs), also known as exosomes, have been already recognized as important mediators of intercellular and extracellular communications in the cardiovascular system^5^. Both proteins and microRNAs (miRNAs) have been found to mediate cellular responses induced by sEVs, including cardiac remodeling and cardiac protection^6, 7, 8, 9^. Despite the interest for sEVs in cardiovascular research, the understanding of the role of endogenous cardiac sEVs in the local microenvironment has been so far hindered by the lack of appropriate research models. Previous *in vitro* and *in vivo* studies have adopted sEVs prepared from either complex and non-cardiac specific samples, such as the human plasma^10^, or cultured cells, which are unable to recapitulate the complexity of the myocardium, making most studies of low translational value ^2^.

Living myocardial slices (LMS) are 300 µm thin layers of ventricular myocardial tissue that can be obtained from human heart of donors and heart failure patients and, also, from different small and large animal species ^11^. LMS not only retain the multi- and hetero-cellular composition, the electrophysiology, and the extracellular matrix of the adult myocardium, but also possess the ability to long term culture while performing physiological functions, the ability to be manipulated in a specific and controlled manner by various stimuli, and the capacity to be studied for cellular and molecular mechanisms ^11–13^. Noteworthy, pathological stimuli known to affect cardiac remodelling and sEVs biology, such as mechanical overload, can be reliably reproduced in LMS ^14–16^.

In this study, we have employed LMS to isolate and characterize sEVs released from human donors and failing hearts. We have complemented the human studies with analyses of rat LMS subjected to different degrees of mechanical load to investigate the effects of cardiac-derived sEVs on the regulation of cardiac contractility and structural and molecular overload-induced myocardial remodelling.

## Methods

Methods are described in detail in the Supplemental Material.

### Ethical statements for the use of human and rat hearts

Human failing hearts were obtained from the NIHR Cardiovascular Biomedical Research Unit at the Royal Brompton and Harefield NHS Foundation Trust and Imperial College London. The study was approved by a UK institutional ethics committee (NRES ethics number for biobank samples: 09/H0504/104 + 5; Biobank approval number: NP001-06-2015 and MED_CT_17_079) and Imperial College London.

Human donor hearts were obtained from the NHS Blood and Transplant INOAR program (IRAS project ID: 189069) approved by the NHS Health Research Authority, in accordance with the Governance Arrangements for Research Ethics Committees. This study is fully compliant with the Standard Operating Procedures for Research Ethics Committees in the UK. Informed consent was obtained from each patient/family involved in this study.

All experiments performed on animals were approved by the UK Home Office, in accordance with the United Kingdom Animals (Scientific Procedures) Act 1986 and Amendment Regulations 2012; killing procedures were performed in accordance with the established guidelines of the European Directive on the protection of animals used for scientific purposes (2010/63/EU). The investigation conforms to the Guide for the Care and Use of Laboratory Animals published by the US National Institutes of Health (NIH Publication No. 85-23, revised 1985) and to the principles outlined in the Declaration of Helsinki (Br Med J 1964; ii: 177).

### Statistical analysis

All data are expressed as mean ± standard error of the mean. Data were analysed using GraphPad Prism 9 software. To assess normality, Shapiro-Wilk’s test was used. For multiple groups comparisons of normally distributed data where unpaired samples were used, such as contractility data in physiological, overloaded and EVs treated LMS at different stretch increments, two-way ANOVA was used, followed by Tukey’s multiple comparison test. For multiple groups comparison of normally distributed data where paired samples were used, such as contractility data in human failing LMS treated with donor sEVs and untreated, multiple paired t-test was used, as control and treated preparation were handled in parallel. To measure sEVs particle size distribution in human and rat samples, the area under the curve for the particle size distribution was calculated, and an unpaired 2-tail t-test performed, using the average concentration, SEM and N number for the respective conditions, which were previously calculated in the area under the curve analysis. For experiments assessing remodelling aspects in physiological, overloaded and EVs treated LMS, one-way ANOVA was used followed by Tukey’s multiple comparison test. Kruskal Wallis test and Dunn’s multiple comparison test were used when data were not normally distributed. 2-tailed student t-test was used to compare the treated versus non-treated groups in iPSC-CMs-only and co-culture experiments.

## Results

### Active force in human failing LMS is significantly lower compared with human donor LMS

All human donor and failing LMS were cultured at a SL of 2.2 µm. To investigate contractility, we performed a force-stretch relationship experiment after 48 hours of culture. As shown in Figure 1 A and B, donor LMS generated a larger force upon stimulation compared with failing LMS; there was no significant change in passive force (Figure 1C).

**Figure 1.**
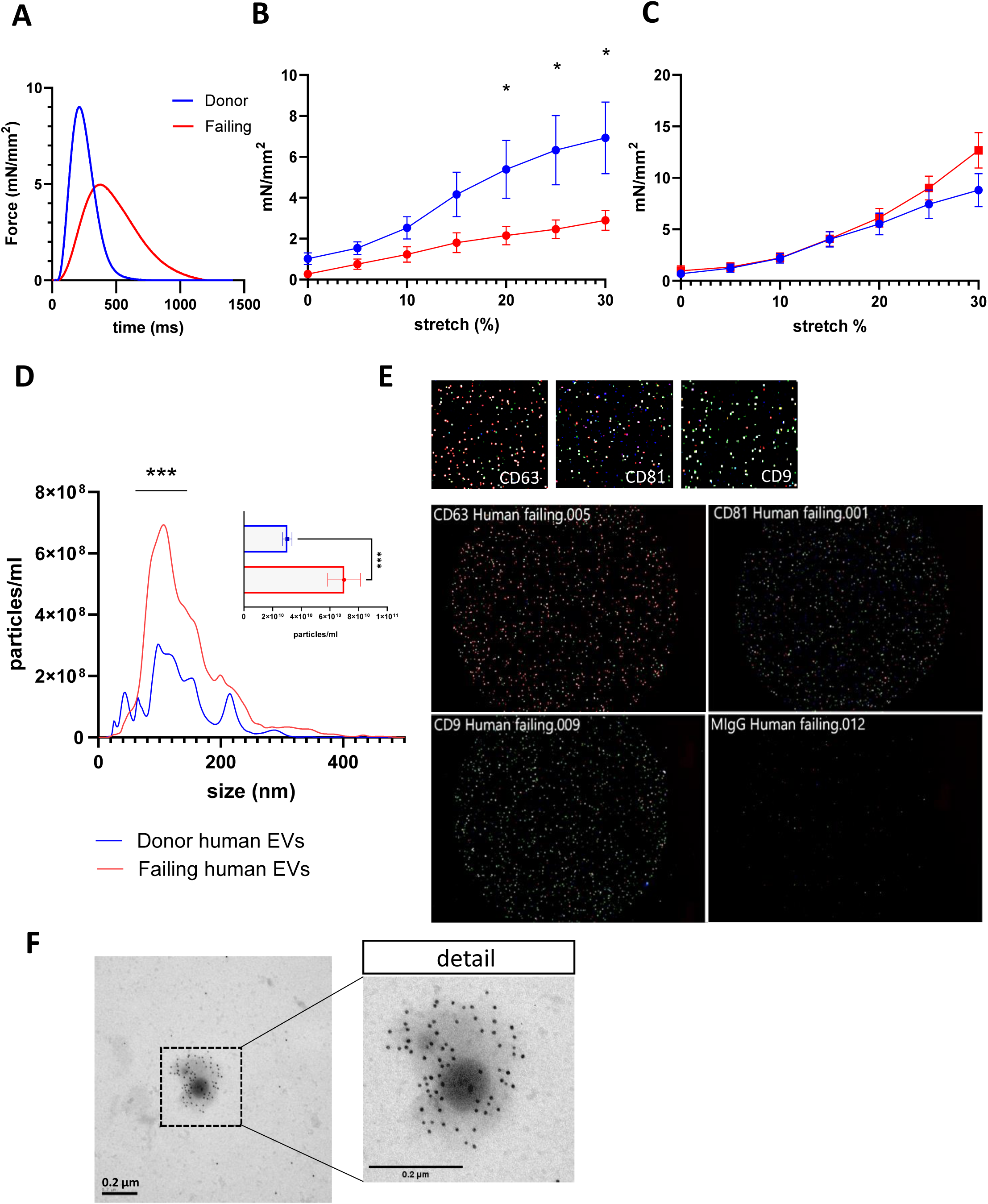
Characterization of human LMS contractility and human LMS-derived sEVs. **A.** Representative traces of produced force of donor and failing LMS at 25% stretch. **B.** Active force and **C.** passive force of human donor and failing hearts after 48 hours culture. N=6 Donors and N=9 Failing. Data are presented as mean+/− SEM. Multiple unpaired t-tests. *P<0.05. **D.** Particle size distribution of failing and donor sEVs. N=3 donor sEVs and N=6 failing sEVs. Unpaired t-test. ***P<0.001. **E.** Nanoview representative image of nanoparticle analysis of failing human sEVs. CD63 (red), CD81 (green), CD9 (blue) and **F.** Transmission electron microscopy with CD63 immunogold labelling of human donor sEVs (left panel) and zoomed in view (right panel).

### Human failing sEVs are more abundant compared to human donor EVs

Cardiac sEVs were extracted from the supernatant of donor and failing LMS culture. The sEV particle-size distribution and concentration were analysed by Nanoparticle Tracking Analysis. The analysis showed a higher concentration of sEVs/ml from the failing heart, compared to the donor heart (Figure 1 D). The sEV identity was confirmed by tetraspanin (CD63, CD81 and CD9) chips for sEV capture and imaging on the ExoView™ R100 imaging platform (NanoView Biosciences). As shown in Figure 1 E, the human sEVs expressed all three tetraspanins ^17^, in different quantities, with CD81 being the most abundant (Supplementary figure 1 B). Transmission electron microscopy with immunogold-labelled antibodies targeting the human CD63 tetraspanin marker further confirmed the successful enrichment of sEVs secreted by human LMS (Figure 1 F).

### Human sEVs released by the donor LMS increase the active force produced by human failing LMS

Given the biological relevance of cardiac sEVs isolated from the human donor LMS, we sought to investigate whether these sEVs affect cardiac contractility of human failing LMS. We observed that failing human LMS cultured with 10^9^ donor sEVs, which corresponded to the number of particles released from an individual human donor LMS after 48 hours, had increased contractility compared with their untreated counterpart (Figure 2 A). In fact, the active force of failing LMS treated with donor sEVs was significantly higher at 15 and 20% stretch compared with non-treated LMS, whilst no change in passive force was observed (Figure 2 B and C).

**Figure 2.**
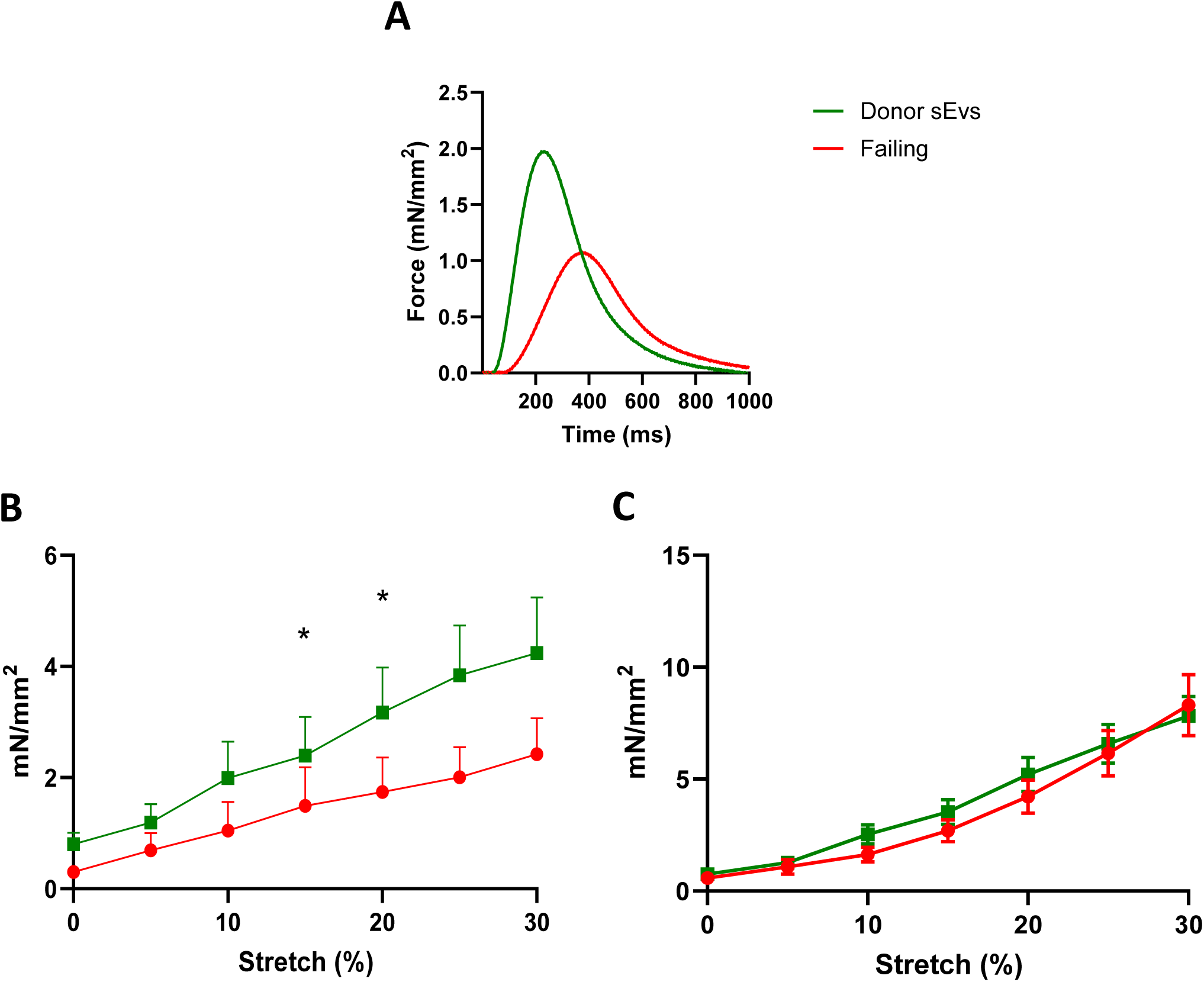
Human donor sEVs significantly increase the active force produced by human failing slices at physiological load. **A.** Representative traces of the force generated by failing slices treated with donor sEVs and control. **B.** Active force and **C.** Passive force of the force-stretch relationship of human failing slices treated with donor sEVs. N=5 Failing, N=5 Failing+Donor sEVs. For each N, the force-stretch measurements of 3-4 LMS were averaged. Data are presented as mean +/− SEM. Multiple paired t-tests. *P<0.05.

### Active force production in rat physiological LMS is higher compared with overloaded LMS

To recreate physiological load and overload, we stretched rat slices uniaxially in order to obtain a SL of 2.2 and 2.4 µm, respectively (Figure 3 A, from ^14^). Following 48 hours of culture, we observed that physiological and overloaded LMS exhibited a different contractile phenotype (Figure 3 B). Figure 3 C shows that physiological LMS generated significantly higher active force from 5 to 25% stretch, compared with the overloaded LMS. The passive force did not change significantly between the two conditions (Figure 3 D).

**Figure 3.**
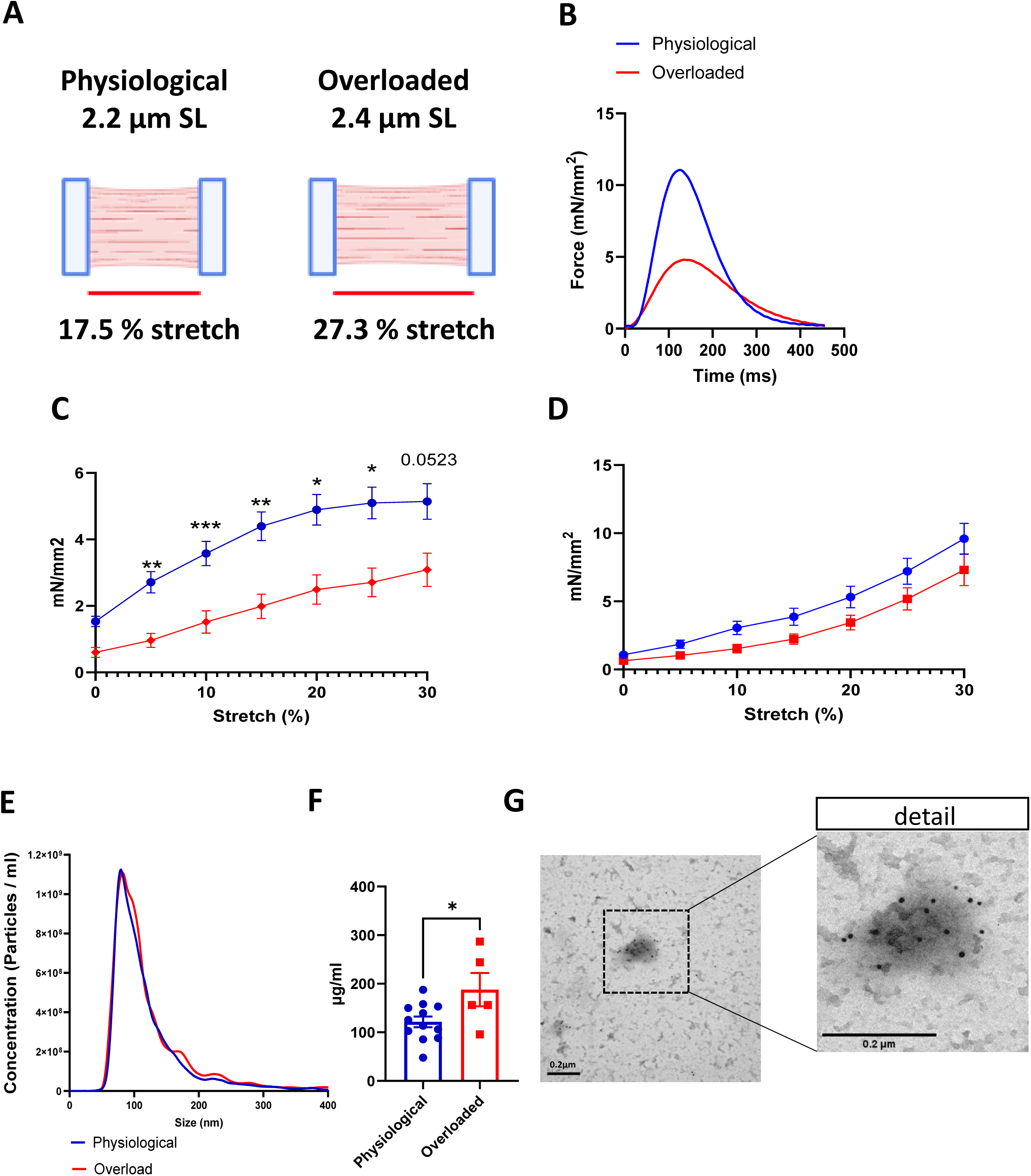
Characterization of rat LMS contractility and rat LMS-derived sEVs. **A.** Representative images of rat slices stretched at 2.2 and 2.4 µm of SL to recapitulate physiological load and overload, respectively (created in BioRender.com). **B.** Representative trace of contractility of physiological and overloaded slices at 25% stretch. **C.** active force and **D.** passive force of rat physiological and overloaded slice after 48 hours culture. Data are presented as mean +/− SEM. N=28 physiological and N=19 overloaded slices. 2-Way-ANOVA analysis with Šídák’s multiple comparisons test. *P<0.05; **P<0.01. **E.** Particle size distribution of physiological and overloaded EVs from rat slices. N=16 physiological EVs and N=8 overloaded EVs. Unpaired t-test. **F.** Quantification of internal protein content of EVs from physiological and overloaded slices from rat slices. Data are presented as mean +/− SEM. Unpaired t-test. *P<0.05. **G.** Transmission electron microscopy with CD63 immunogold labelling rat physiological EVs (left panel) and zoomed in view (right panel).

### Rat overloaded LMS secrete an equal amount of sEVs to physiological LMS but the sEVs contain more proteins

A particle-size distribution analysis revealed a similar concentration of sEVs released from overloaded LMS compared to physiological LMS (Figure 3 E and supplementary figure 1 A). However, measurements of internal protein content of sEVs revealed a higher amount of protein in the sEVs from overloaded LMS, when compared to physiological LMS (Figure 3 F). Electron microscopy with CD63 immunogold labelling confirms the presence of rat sEVs (Figure 3 G). Given the more immediate availability of rat hearts, the following functional, structural, and molecular studies were performed on rat LMS and rat LMS-derived sEVs.

### Physiological cardiac sEVs increase the active force and decrease the passive force of physiological LMS

Both physiological and overloaded sEVs were applied on rat physiological LMS to assess whether sEVs would mediate changes in contractility on healthy tissue (Figure 4 A). Force-stretch relationship of physiological LMS treated with either PBS vehicle, physiological sEVs or overloaded sEVs revealed that physiological sEVs increase the active force of physiological LMS at 25% and 30% stretch (Figure 4 B) and decrease the passive force from 0 to 20% stretch (Figure 4 C). However, the application of overloaded sEVs on physiological LMS had no significant effect on the production of both active and passive force, compared to untreated physiological LMS (Figure 4 B and C).

**Figure 4.**
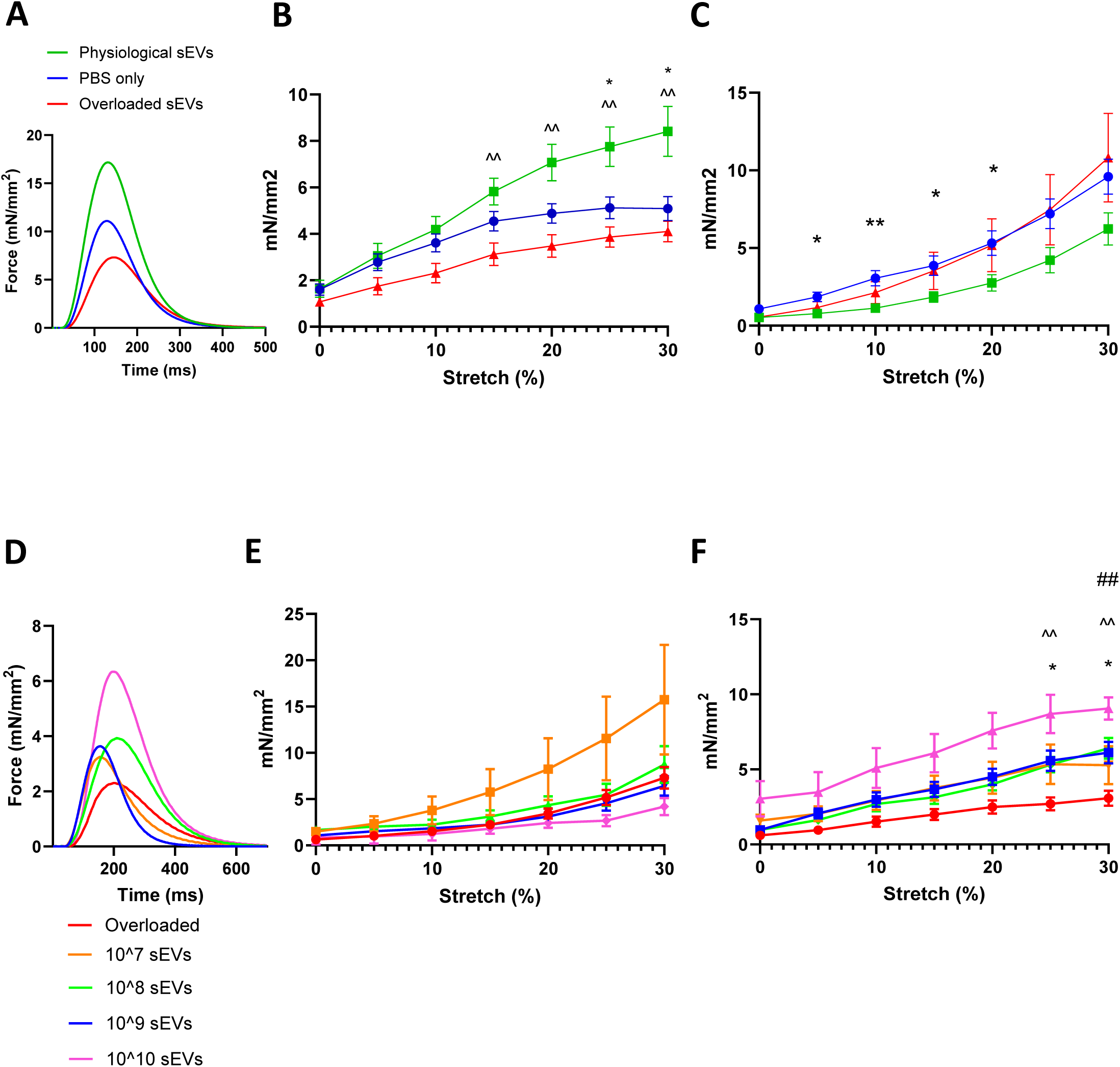
The effect of cardiac sEVs on the force generation of rat living myocardial slices. **A.** Representative traces of physiological slices treated with physiological sEVs, overloaded sEVs and control. **B.** Active force and **C.** Passive force of the force stretch relationship of rat physiological slices treated with physiological sEVs, overloaded sEVs or control (PBS only). N=33 PBS treated LMS, N=11 Physiological sEVs LMS, N=14 Overloaded sEVs LMS. Data are presented as mean +/− SEM. 2-Way ANOVA with Tukey’s multiple comparisons test. *P<0.05 and **P<0.01 for PBS only vs physiological sEVs, ^^=P<0.01 Physiological sEVs vs overloaded sEVs. **D.** Representative traces of dose-response experiments. **E.** Active force and **F.** Passive force of the force stretch relationship of rat overloaded slices treated with 10^7, 10^8, 10^9, 10^10 sEVs per slices, or control (overloaded). N=19 overloaded, N= 7 10^7, N= 11 10^8, N=16 10^9, N=3 10^10. Data are presented as mean +/− SEM. 2-Way ANOVA with Tukey’s multiple comparisons test. *=P<0.05 overloaded vs 10^9 sEVs, ^^=P<0.01 overloaded vs 10^8 sEVs, ##=P<0.01 overloaded vs 10^10 sEVs.

### Physiological cardiac sEVs increase the active force of overloaded LMS in a dose-response manner

To assess the functional effect of rat physiological sEVs on the overloaded LMS, concentrations ranging from 10^7^ to 10^10^ were tested (Figure 4 D). We observed that the minimum amount of sEVs per LMS required to achieve a significant increase in active force was 1×10^8^ sEVs (Figure 4 E). However, no significant difference was observed in the passive force of overloaded rat LMS treated with physiological sEVs (Figure 4 F). The following structural and molecular experiments to assess cardiac remodelling were performed using 1×10^8^ sEVs.

### Application of physiological cardiac sEVs does not affect the size of cardiomyocytes in overloaded LMS

In a condition of volume overload, eccentric hypertrophy develops, where the cardiomyocytes elongate, and sarcomeres are added in series ^18^. In our model of uniaxial mechanical overload, we saw a significant elongation of the cardiomyocytes, compared to the physiological condition (Supplementary figure 2 A). This resulted in an overall increase of the cardiomyocytes area, whilst no difference was observed in the width (Supplementary Figure 2 B and C). The application of physiological sEVs on overloaded slices did not change cell size (Supplementary Figure 2 D).

### Application of physiological cardiac sEVs attenuates the transcriptional expression of profibrotic genes in overloaded LMS

The development of fibrosis following mechanical overload is an important determinant of cardiac contractility^19^. Production and deposition of collagens in the ECM and proliferation of stromal cells are crucial factors in the identification of cardiac fibrosis. Therefore, we first examined the amount of Collagen I and Vimentin deposition through immunostaining (Figure 5 A); no difference was observed between physiological and overloaded LMS (Figure 5 B and C, respectively). The expression of Collagen I was also studied at protein level, along with the protein α-smooth muscle actinin (α-SMA), to assess fibroblasts activation. No significant change was observed in the expression of Collagen I (Figure 5 D) and α-SMA

**Figure 5.**
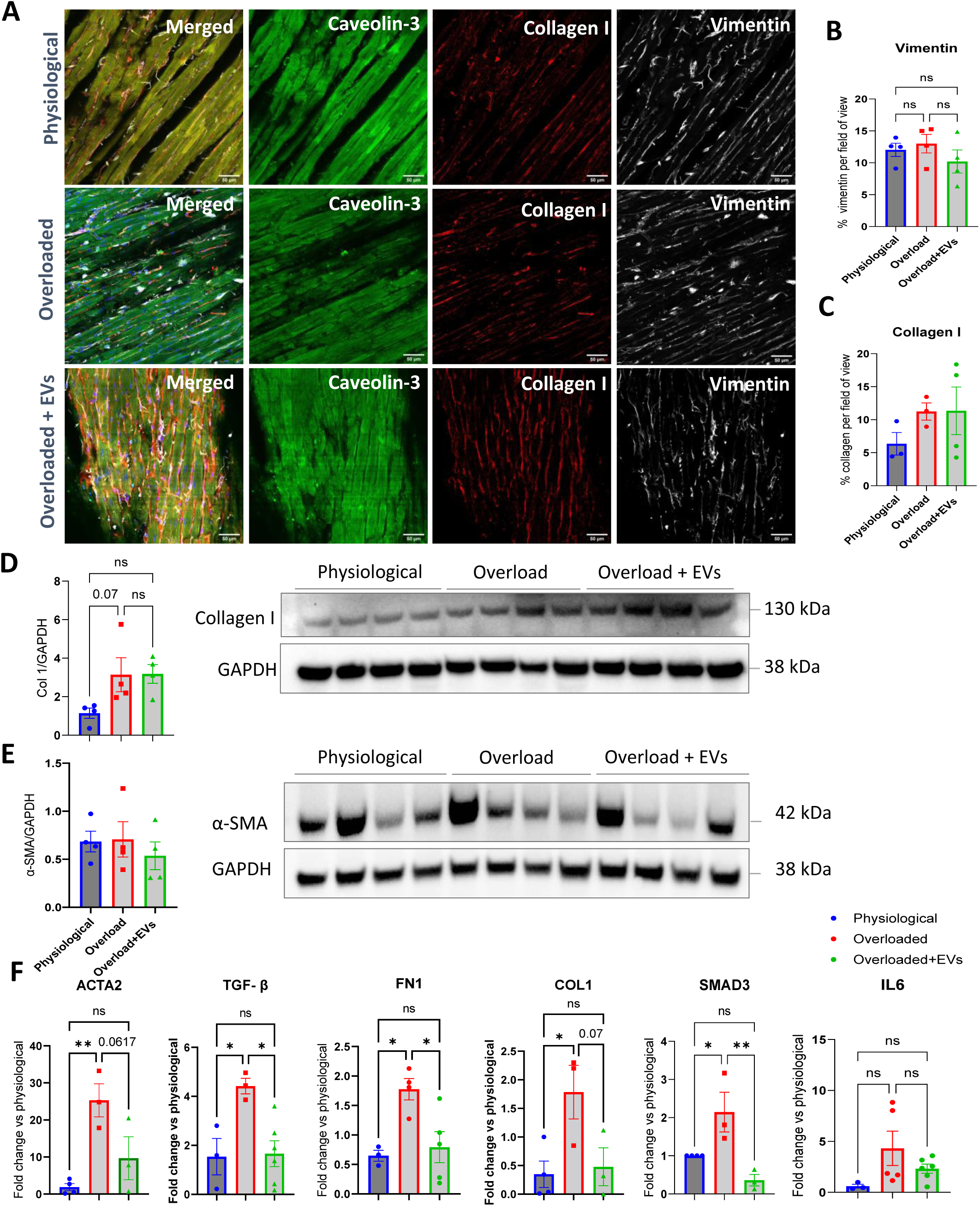
The effect of physiological sEVs on the fibrosis of overloaded slices. Figure 5. **A.** Representative images of physiological, overloaded and overloaded + sEVs slices stained for Caveolin 3, Vimentin and Collagen I. Scale bar 50 µm. Quantification of immunofluorescence staining of **B.** Vimentin and **C.** Collagen I. Data are presented as mean +/− SEM. One-Way ANOVA with Tukey’s multiple comparisons test.. **D.** Western Blot quantification of Collagen I expression in physiological, overloaded and overloaded slices+sEVs. One-Way ANOVA with Tukey’s multiple comparisons test. **E.** Western Blot quantification of α-SMA expression in physiological, overloaded and overloaded + sEVs slices. Data are presented as mean +/− SEM. One-Way ANOVA with Tukey’s multiple comparisons test. **F.** mRNA quantification of the expression of major pro-fibrotic markers in physiological, overloaded and overloaded + sEVs slices. Data are presented as mean +/− SEM. One-Way ANOVA with Tukey’s multiple comparisons test. =P<0.05, **p<0.01.

(Figure 5 E) between physiological and overloaded slices. Likewise, the application of physiological sEVs on overloaded slices did not have any effect on fibrosis development at either structural or protein level. However, when we analysed expression of some of the fibrosis-related genes by q-RT-PCR, we observed increased levels of α-SMA, Fibronectin, TGF-β, Collagen I and SMAD3 in the overloaded LMS, compared with physiological. Application of physiological sEVs on overloaded slices prevented/reversed the activation of Fibronectin, TGF-β and SMAD3 (Figure 5 F).

### Application of physiological cardiac sEVs reduces cell death in overloaded LMS

To assess cell death, another possible determinant of reduced cardiac contractility ^20^, we first measured the level of cleaved PARP-1. PARP-1 is cleaved when it gets activated during apoptosis ^21^. Cleaved PARP-1 was increased in overloaded LMS compared with physiological LMS. Moreover, upon application of physiological sEVs, the cleavage of PARP-1 was reduced to almost physiological levels (Figure 6 A). Bcl-2 is a potent pro-survival protein. Whilst its expression was maintained at basal level in physiological and overloaded slices, the level of Bcl-2 increased following application of sEVs on overloaded LMS. These data suggest that sEVs activate pro-survival pathways in the slices subjected to increased mechanical load (Figure 6 B).

**Figure 6.**
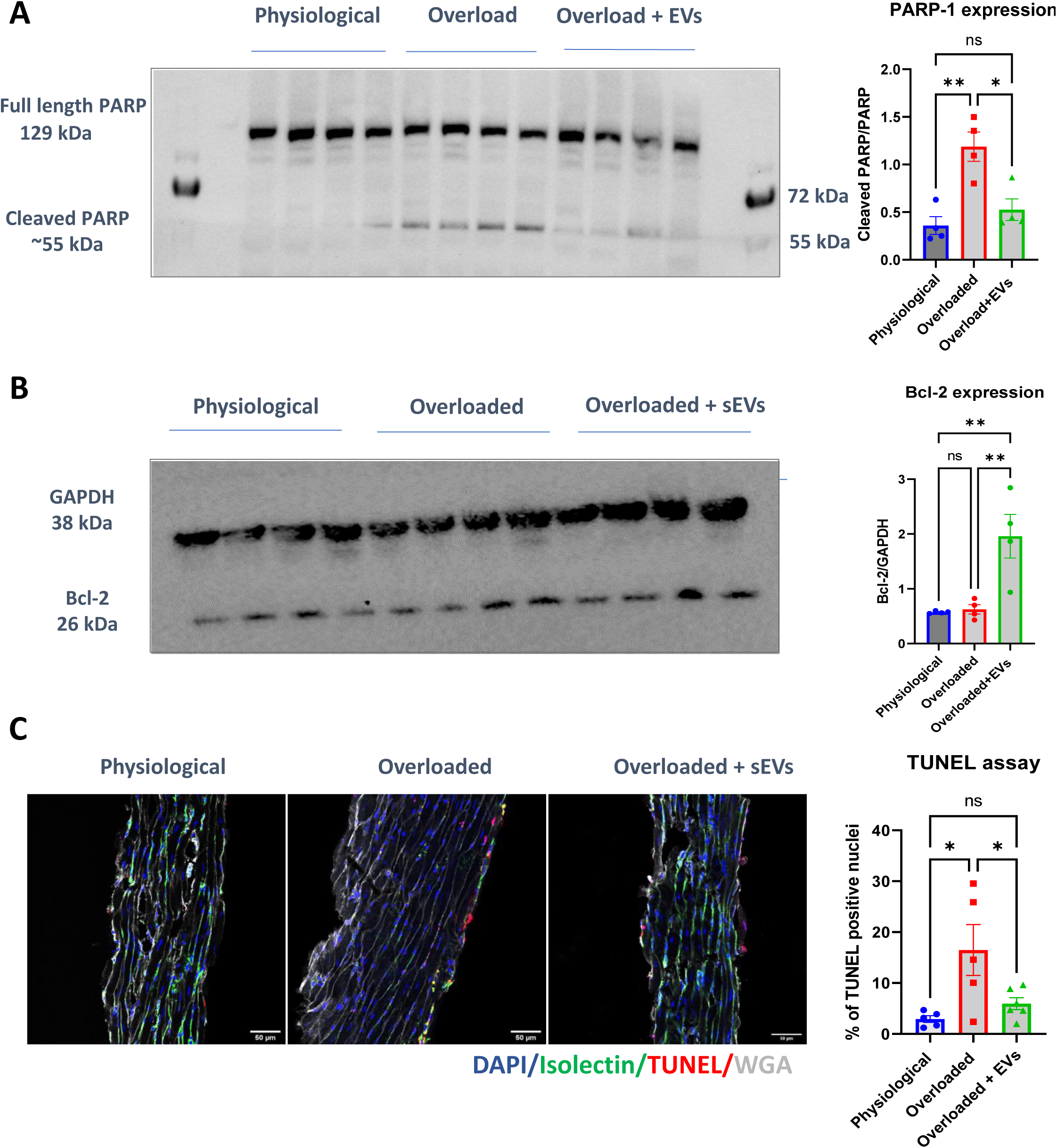
The effect of physiological sEVs on the apoptosis of overloaded slices. **A.** Western Blot quantification of PARP-1 expression in physiological, overloaded and overloaded + sEVs slices. One-Way ANOVA with Tukey’s multiple comparisons test. *P<0.05, **P<0.01. **B.** Western Blot quantification of Bcl-2 expression in physiological, overloaded and overloaded + sEVs slices. One-Way ANOVA with Tukey’s multiple comparisons test. **P<0.01. **C.** Quantification of immunofluorescence staining of TUNEL assay in physiological, overloaded and overloaded slices+ sEVs. Slices were stained with DAPI, Isolectin B4, TUNEL and WGA. One-Way ANOVA with Tukey’s multiple comparisons test. *=P<0.05. Scale bar 50 µm.

Cell death in the LMS was further investigated by TUNEL staining. Transversal LMS cryosections showed increased TUNEL positive nuclei mostly on the surface of overloaded LMS, compared with physiological LMS. Application of physiological sEVs on overloaded LMS reduced the abundance of TUNEL positive nuclei to almost physiological levels (Figure 6 C).

### Application of physiological cardiac sEVs reduces microvascular rarefaction in overloaded LMS

Heart failure is associated with microvascular rarefaction^22,23^. In our study, Isolectin B4 staining confirmed reduced microvascular density in the overloaded LMS compared with physiological slices. Application of physiological sEVs in overloaded slices partially prevented this phenomenon (Figure 7 A) and increased the mRNA expression of VEGF-A, a prototypical proangiogenic and angiocrine factor (Figure 7 B).

**Figure 7.**
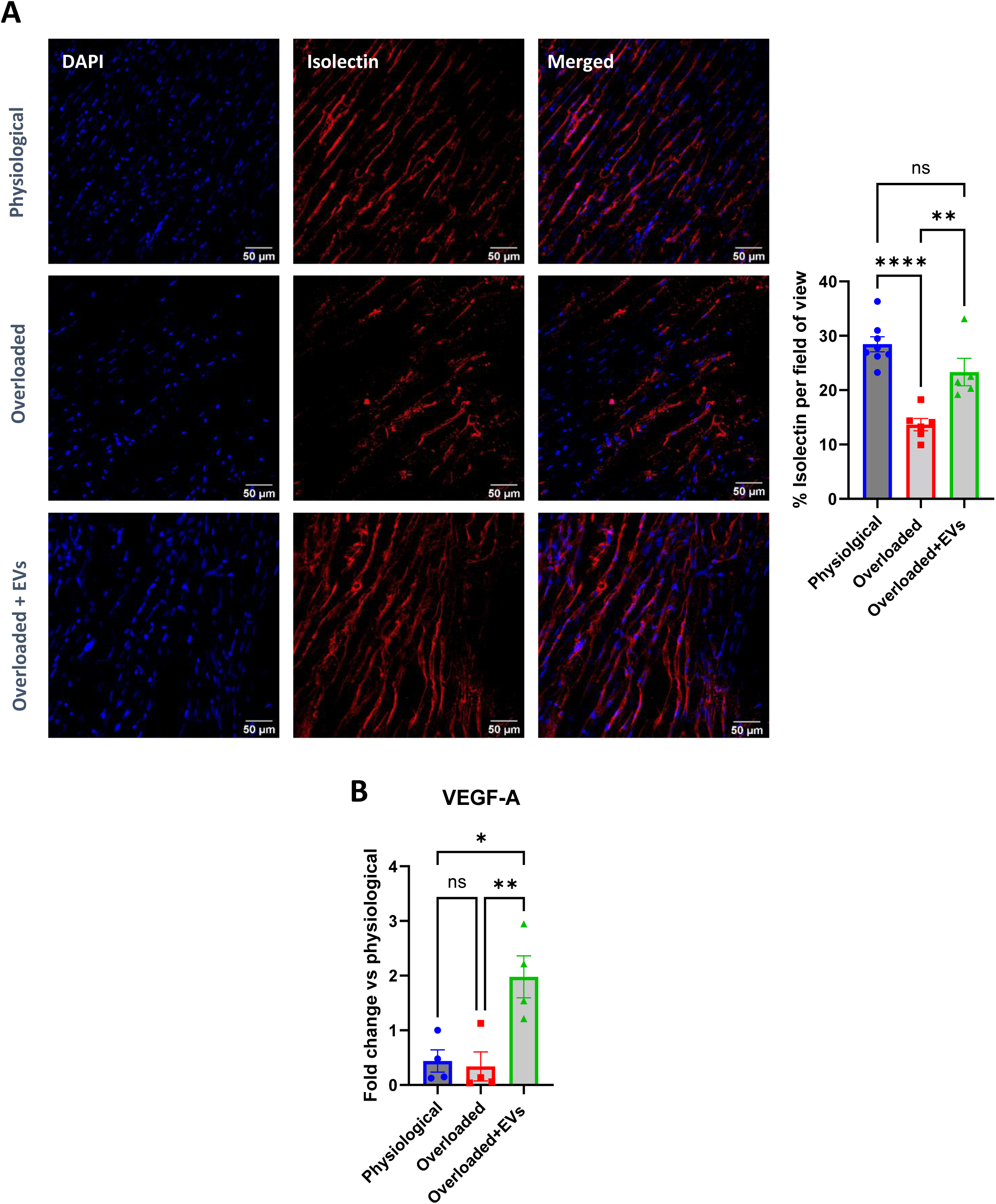
The effect of physiological sEVs on the blood vessel rarefaction in overloaded slices. **A.** Quantification of Isolectin B4 immunostaining in physiological, overloaded and overloaded slices + sEVs. Scale bar 50 µm. Data are presented as mean +/− SEM. One-Way ANOVA with Tukey’s multiple comparisons test. **P<0.01, ****P<0.0001. **B.** Quantification of VEGF gene expression by qPCR in physiological, overloaded and overloaded + sEVs slices. Data are presented as mean +/− SEM. One-Way ANOVA with Tukey’s multiple comparisons test. over physiological slices. *P<0.05. Data are expressed as fold change expression.

**Figure 8.**
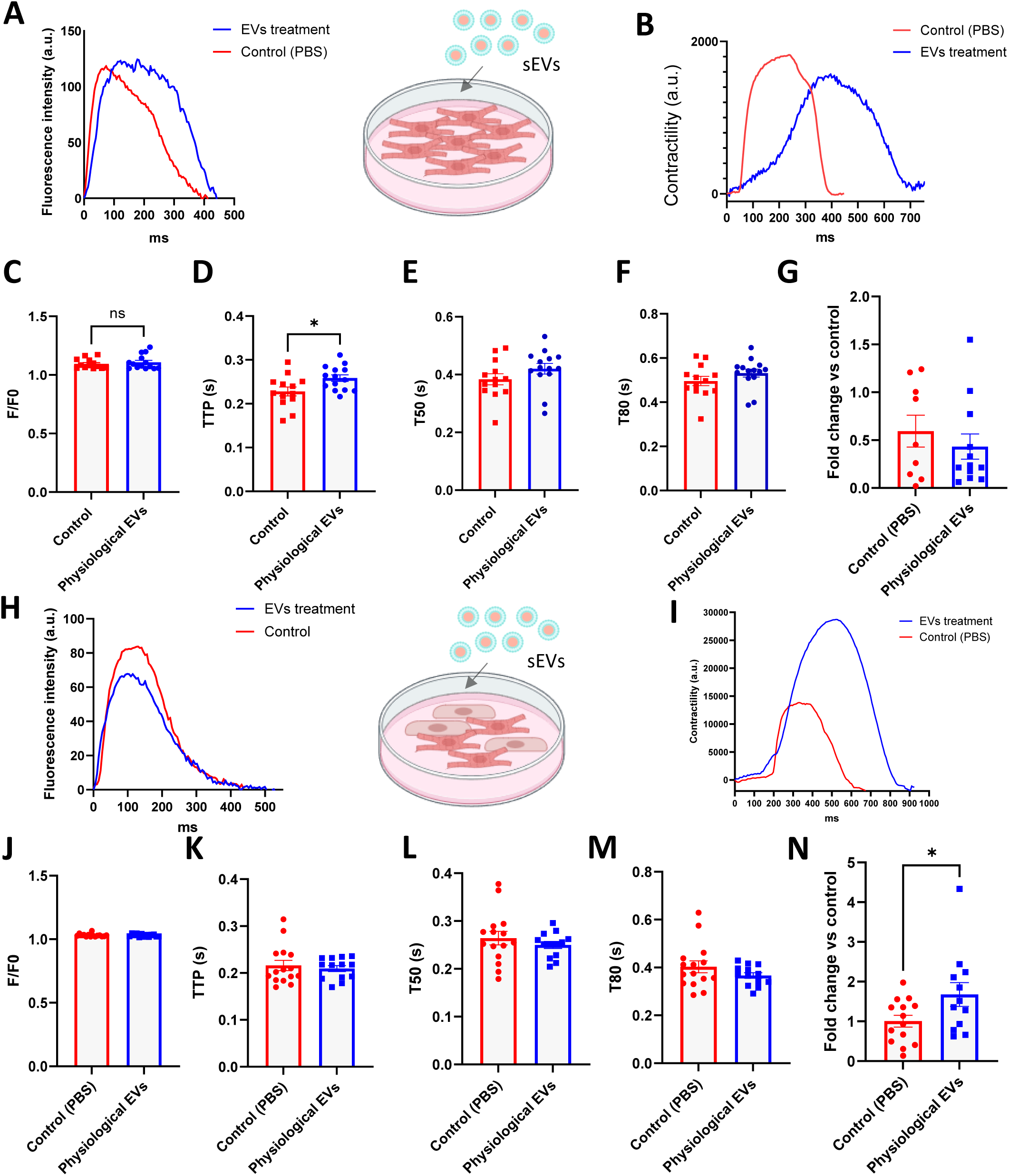
The effect of physiological sEVs on iPSC-CMs and on iPSC-CMs cultured with microvasculature endothelial cells. **A.** Representative traces of calcium transients in iPSC-CMs treated with physiological sEVs and control. **B.** Representative traces of contractility in iPSC-CMs treated with physiological sEVs and control **C.** Amplitude **D.** Time to peak **E.** Time to 50% decay and **F.** Time to 80% decay of calcium transients and **G.** amplitude of contractility in iPSC-CMs treated with rat physiological sEVs or control. Unpaired t-test. *=P<0.05. **H.** Representative traces of calcium transient amplitude in iPSC-CMs and microvasculature ECs co-culture treated with physiological sEVs and control. **I.** Representative traces of contractility in iPSC-CMs and microvasculature ECs co-culture treated with physiological sEVs and control **J.** Amplitude **K.** Time to peak **L.** Time to 50% decay and **M.** Time to 80% decay of calcium transients and **N.** amplitude of contractility in iPSC-CMs co-cultured with microvasculature endothelial cells treated with rat physiological sEVs or control. Data are presented as mean +/− SEM. Unpaired t-test. *P<0.05.

### Exogenous cardiac sEVs preferentially localize around the nuclei of non-cardiomyocytes in the recipient LMS

We first performed sEV incorporation studies at 1, 24 and 48 hours using ExoGlow-stained sEVs to understand the time course of sEV uptake. We observed the presence of sEVs on the surface of LMS at 1- and 24-hours, but this was reduced at 48-hours (Supplementary Figure 3 A), suggesting that after 48 hours of incubation, sEVs are mostly internalised by the LMS. To assess the biodistribution of sEVs, we performed experiments using the 24-hours timepoint, to quantify the presence of sEVs on the cells surface at their maximum visibility. We observed that sEVs were preferentially localized around the nuclei of non-cardiomyocytes: 19.3% of the total nuclei of vimentin-positive cells, and only 5.76% of α-actinin-positive cells were co-localised with sEVs (Supplementary Figure 3 B).

### Physiological cardiac sEVs contain higher level of microRNA-378a-3p and −23a-3p in comparison with pathological sEVs

To investigate differences in miRNAs cargo between physiological and overloaded rat sEVs, we performed a miRNA array to assess the expression of 372 miRNAs which are conserved between rat and human species. We found two miRNAs (miR), miR-378a-3p and hsa-miR-23a-3p, to have higher expression in physiological compared with overloaded sEVs. By contrast, miR-16-5p and miR-346, were significantly increased in the overloaded sEVs (Supplementary Figure 4 A and B). We focussed on the upregulated miRNAs in physiological sEVs, as these could directly repress the expression of their target genes in overloaded LMS. miRNA qPCR analysis confirmed the upregulation of miR-378a-3p and miR-23a-3p in physiological sEVs compared with overloaded sEVs (Supplementary Figure 4 C and D).

### Bioinformatics analysis reveals potential mRNA targets of miR-378a-3p and miR-23a-3p that are increased in cardiomyocytes extracted from overloaded LMS

We performed a bioinformatics analysis to find potential target genes for miR-378a-3p and hsa-miR-23a-3p. Predicted targets of miR-378a-3p and hsa-miR-23a-3p were cross-referenced with genes that were significantly differentially expressed in cardiomyocytes isolated from overloaded LMS, compared to physiological (Supplementary Figure 5 A). Six target genes, Gadd45g, Slc4a11, Xirp2, Dusp13, Usp13 and Triadin, were identified, which were overexpressed in overloaded cardiomyocytes and regulated by miR-378a-3p and hsa-miR-23a-3p (Supplementary Figure 5 B and C). Pathway analyses performed on these genes revealed their possible involvement in cardiac contractility and apoptosis (Supplementary Figure 5 D).

### Physiological cardiac sEVs do not regulate the contractility of human cardiomyocytes monocultures

Given the interplay between different cardiac cells in the LMS, it is challenging to assess whether cardiomyocytes are directly affected by sEVs to increase cardiac contractility. To address this issue, we employed human iPSC-CMs that express the calcium indicator GCaMP6f ^24^, to simultaneously assess the impact of cardiac sEVs on calcium handling and cardiomyocyte contractility (Figure 10 A and B). iPSC-CMs were seeded on 7 mm diameter MatTek dishes at a density of 80,000 cells per dish for 48 hours and treated with 10^9^ sEVs or PBS control. Calcium handling and contractility were assessed and compared between sEVs-treated and untreated (PBS only) iPSC-CMs. There was no significant difference in the amplitude of calcium transients between sEV-treated iPSC-CMs and controls (Figure 10 C). Nevertheless, the iPSC-CMs time to peak of the calcium transients was increased after sEV-treatment (Figure 10 D). The time to 50% and 80% decay remained unchanged between the 2 conditions (Figure 10 E and F). Likewise, the amplitude of iPSC-CM contractility was not different in sEV-treated and control iPSC-CMs (Figure 10 G).

### Physiological cardiac sEVs increase contractility of IPSC-CM co-cultured with endothelial cells

Given the increased VEGF-A and vascular response observed in overloaded LMS treated with physiological cardiac sEVs (Figure 7 A and B), and the prevalent non-CM distribution of exogenous sEVs in recipient LMS (supplementary Figure 3 B), we hypothesised the importance of endothelial cells for the CM response to sEVs. We therefore co-cultured iPSC-CMs with human coronary microvascular endothelial cells (MVECs) before sEVs application (Figure 10 H and I). Physiological cardiac sEVs did not affect calcium handling parameters compared to control (Figure 10 J-M). However, cardiac sEV increased the contractility of iPSC-CM co-cultured with MVEC (Figure 10 N).

## Discussion

We report the successful isolation and characterisation of sEVs secreted from the ventricular myocardium of human failing and donor healthy hearts and from rat LMS cultured under physiological and overloaded conditions. Our protocols open unprecedented possibilities for the characterization of the cardiac sEVs in the cardiac microenvironment.

This study has demonstrated that cardiac sEVs released from the human donor hearts increase the contractility of failing LMS. Moreover, rat physiological sEVs increase contractility of both physiological and overloaded LMS, at a minimum concentration of 10^8^ particles/LMS. In contrast, sEVs released from the rat overloaded LMS had lost the capacity to regulate contractility.

We have showed that overload reduces LMS contractility, increases the expression of profibrotic genes, reduces cell survival and induces microvascular rarefaction, simulating the feature of failing myocardium. Importantly, these pathogenic features are repressed by stimulating the LMS with rat physiological cardiac sEVs, thus suggesting that sEVs released from healthy hearts contribute to preventing and possibly reversing, pathological cardiac remodelling.

HF with reduced ejection fraction (HFrEF) is characterised by lower muscle contractility. In clinical cases of HF, there is a downward shift in the Frank-Starling curve, which correlates with the inability of the heart to work effectively ^28^. LMS are an ideal *ex vivo* platform to study myocardial contractility because they retain the electrophysiology and multicellularity of the adult myocardium ^29^. Here we show that human failing LMS shifted the Frank Starling curve downwards compared with donor LMS, in accordance with what is observed clinically ^30,31^.

By applying different degrees of mechanical load on rat LMS, we were able to induce a distinct contractile phenotype, similar to what was observed in the human slices. This finding has been previously reported by us in ^14,32^. For instance, we experimentally determined the best SL to culture LMS for 24 hours, and found that slices cultured at 2.2 µm maintained the same viability, gene expression and minimal change in force compared to fresh slices, where contractility was measured immediately after slicing ^14^. We reported a significant difference in the amplitude of maximum contractility between rat slices cultured at 2.2 and 2.4 µm after 48 hours. The significant decrease in force produced by the overloaded slices was associated with an increase in Collagen I deposition and the overexpression of major profibrotic genes ^32^. While 48-hour culture might not be sufficient to induce a complete remodelling of the LMS *in vitro,* the rat overloaded LMS recapitulate several features of human failing myocardium.

Native sEVs are considered for their potential to regulate cardiac homeostasis. This is in line with the knowledge that sEVs isolated from specific cardiac cell types contain cargoes that can confer cardioprotection in a damaged heart ^33^; however, the effects of cardiac sEVs released from the myocardial tissue remain untested. In this study, we demonstrate that LMS release sEVs; we show the ability of healthy sEVs derived from the human and rat hearts to restore contractility and partially reverse/prevent remodelling after cardiac damage ^35–38^.

In our study, despite observing a significant difference in sEV release between human failing and donor LMS, we showed that there was no difference in the concentration of sEVs released by either rat overloaded or physiological LMS after 48 hours culture. However, overloaded LMS-derived sEVs did not significantly affect the production of either active or passive force in physiological slices, in clear contrast with physiological sEVs, suggesting that the formulation of the sEVs derived from physiological and pathological is different. In line with this, we recorded different levels of total protein content between overloaded and physiological sEVs.

Several studies have shown that sEV actions are mediated by miRNA transfer ^39,40^. Therefore, the beneficial effects of cardiac sEVs may be mediated by miRNA transfer. In line with this hypothesis, we found increased expression of miR-378a-3p and hsa-miR-23a-3p in the physiological cardiac sEVs when compared with the sEV from rat overloaded LMS. These miRNAs have been previously shown to repress cardiac hypertrophy ^41^ and apoptosis ^42^ and to promote angiogenesis ^45^. A limitation of our miRNA array approach is that it contained a limited number of miRNAs selected for their conservation between humans and rats. Alternative miRNA candidates might emerge in future studies based on RNA-sequencing approaches.

A miRNA canonical mechanism of action is to repress the expression of a pool of targeted genes acting at post-transcriptional level by binding to the 3’UTR of mRNAs ^46^. In line with this, bioinformatic target prediction revealed that potential target genes of miR-378a-3p and hsa-miR-23a-3p were involved in the regulation of cardiac contractility and apoptosis of cardiomyocytes.

Excessive mechanical preload of the heart results in the development of eccentric hypertrophy, where cardiomyocytes elongate through the addition of sarcomeres in series ^47^. In our model of mechanical overload, we observed a significant elongation of cardiomyocytes compared to the physiological counterpart. The addition of sarcomeres in series leads to a decrease in force being generated by the cardiomyocytes, as to reach an optimal SL, longer cardiomyocytes may need to be stretched more, compared to shorter cardiomyocytes containing less sarcomeres ^48^.

In our study, we found that after 48 hours of culture under overloaded conditions, the process of fibrosis activation is only initiated at transcriptional level, with no significant changes at protein and structural level. However, physiological sEVs were able to inhibit the overexpression of the major pro-fibrotic genes in overloaded slices. A previous study on fibrosis conducted on LMS showed that, although there was a significantly higher Collagen I deposition in overloaded slices after 48 hours compared with the physiological controls, the difference in the amount of stromal cells proliferation on the slices was unchanged^15^. Moreover, the changes in Collagen I were only seen with immunofluorescence staining and could not be detected by western blot analysis.

The cleavage of PARP-1 is a hallmark of cell death ^49^. In our study, we observed a significant increase in cleaved PARP-1 in the overloaded slices compared with the physiological LMS. TUNEL staining revealed that cell death was prevalently localized on the surface of the myocardial slices. During LMS preparation, the blade cutting along the surface of the tissue damages ∼3% of the total LMS cells, and only on the surface of the slice, leaving the rest of the preparation intact ^11^. We speculate that surviving cells on the LMS surface are more fragile and sensitive to the detrimental actions of overload. Physiological sEVs were able to inhibit the process of cell death on overloaded slices. However, because changes in cell death were limited to the LMS surface, is unlikely that prevention of cell death by the healthy sEVs is the only contributor to the increase in contractility produced by these sEVs.

Microvessels rarefaction is a hallmark of cardiac remodelling and reportedly contributes to HF with preserved ejection fraction (HFpEF)^50^. Microvessels are tasked not only to constantly deliver oxygenated blood and nutrients to cardiomyocytes, but also act as a source of paracrine regulatory molecules that help in the modulation of cardiac contractility ^51^. In the overloaded slices, we observed a significant loss in microvessels. It is worth noting that vessels are not perfused in LMS, therefore regulation of cardiomyocyte contractility is likely to occur through direct contact and/or paracrine interactions. VEGF was maintained at basal levels in physiological and overloaded conditions but, upon application of sEVs, these levels raised significantly, suggesting that sEVs may contain cargoes that promote the production of VEGF. We found that most of the sEVs are taken up by the non-cardiomyocytes. This could indicate an indirect effect on contractility, mediated by other cell types other than CMs, in particular ECs.

To confirm whether sEVs have a direct effect on CM contractility, we analysed calcium handling and contractility of IPSC-CMs monoculture. Physiological sEVs did not affect the contractility of iPSC-CMs nor the amplitude of calcium transients, suggesting that physiological sEVs may act on other cell types. Given the effect of physiological sEVs on LMS microvessels, we co-cultured iPSC-CMs with MVECs. In the co-culture experiments, despite no change in the calcium handling parameters, we observed a significant increase in contractility when physiological sEVs were applied. This suggests that changes in IPSC-CM contractility are: i) mediated by the effect of sEVs on ECs, and ii) due to changes in myofilament sensitivity to calcium. There are few limitations of these cellular experiments: although iPSC-CMs lines have been extensively used in cardiovascular research to recapitulate disease models, these cells are still very different from adult CMs, due to their relatively immature phenotype, and therefore it might not be a fully representative model to describe the changes in contractility that occur in the LMS ^52^. Moreover, in our study mechanical load is a major determinant of remodelling, but overload could not be recreated in iPSC-CMs.

If these results are extrapolated to the LMS model, whereby sEVs had a significant effect on contractility, preserved microvessel density and increased VEGF production, they may suggest that the beneficial effects of sEVs are indirect and mediated, at last in part, by ECs. It could be speculated that once sEVs (and their associated miRNAs) are taken up by ECs, they would release factors (such as VEGF) that improve CM contractility ^53^. As example, miR-378a-3p was already shown to indirectly increase VEGF-A in skeletal muscles ^54^.

## Conclusions

Myocardial-specific sEVs from healthy human and rat hearts restore contractility and reverse/prevent remodelling of failing myocardium. The effects on contractility could be mediated by inhibition of cell death, inhibition of pro-fibrotic gene pathways or by preservation of blood vessels, or a combination of them, via indirect heterocellular interactions. The miRNA signature of the healthy sEVs may contribute to explain these effects. Our findings support the importance of sEVs in physiology and disease, and their potential utilisation in the treatment of cardiovascular disease.

## Data Availability

All the data will be available on request

## Acknowledgements

We would like to thank Steve Rothery for the support with confocal microscopy, Fatemeh Kermani for the gift of MVECs and Danya Agha-Jaffar for the support with the human tissue. We are grateful to the British Heart Foundation for financial support (FS/19/57/34894).

## Bibliography

1. Katz AM. Maladaptive growth in the failing heart: the cardiomyopathy of overload. Cardiovasc Drugs Ther. 2002;16:245–249.

2. Martins-Marques T, Hausenloy DJ, Sluijter JPG, Leybaert L, Girao H. Intercellular Communication in the Heart: Therapeutic Opportunities for Cardiac Ischemia. Trends Mol Med. 2021;27:248–262.

3. Tirziu D, Giordano FJ, Simons M. Cell Communications in the Heart. Circulation. 2010;122:928–937.

4. Jeppesen DK, Zhang Q, Franklin JL, Coffey RJ. Extracellular vesicles and nanoparticles: emerging complexities. Trends Cell Biol. 2023;33:667–681.

5. Davidson SM, Boulanger CM, Aikawa E, Badimon L, Barile L, Binder CJ, Brisson A, Buzas E, Emanueli C, Jansen F, Katsur M, Lacroix R, Lim SK, Mackman N, Mayr M, Menasché P, Nieuwland R, Sahoo S, Takov K, Thum T, Vader P, Wauben MHM, Witwer K, Sluijter JPG. Methods for the identification and characterization of extracellular vesicles in cardiovascular studies: from exosomes to microvesicles. Cardiovasc Res. 2023;119:45–63.

6. Bellin G, Gardin C, Ferroni L, Chachques J, Rogante M, Mitrečić D, Ferrari R, Zavan B. Exosome in Cardiovascular Diseases: A Complex World Full of Hope. Cells. 2019;8:166.

7. Bang C, Batkai S, Dangwal S, Gupta SK, Foinquinos A, Holzmann A, Just A, Remke J, Zimmer K, Zeug A, Ponimaskin E, Schmiedl A, Yin X, Mayr M, Halder R, Fischer A, Engelhardt S, Wei Y, Schober A, Fiedler J, Thum T. Cardiac fibroblast–derived microRNA passenger strand-enriched exosomes mediate cardiomyocyte hypertrophy. Journal of Clinical Investigation. 2014;124:2136–2146.

8. Zheng D, Huo M, Li B, Wang W, Piao H, Wang Y, Zhu Z, Li D, Wang T, Liu K. The Role of Exosomes and Exosomal MicroRNA in Cardiovascular Disease. Front Cell Dev Biol. 2021;8.

9. Vicencio JM, Yellon DM, Sivaraman V, Das D, Boi-Doku C, Arjun S, Zheng Y, Riquelme JA, Kearney J, Sharma V, Multhoff G, Hall AR, Davidson SM. Plasma Exosomes Protect the Myocardium From Ischemia-Reperfusion Injury. J Am Coll Cardiol. 2015;65:1525–1536.

10. Pant S, Hilton H, Burczynski ME. The multifaceted exosome: Biogenesis, role in normal and aberrant cellular function, and frontiers for pharmacological and biomarker opportunities. Biochem Pharmacol. 2012;83:1484–1494.

11. Watson SA, Scigliano M, Bardi I, Ascione R, Terracciano CM, Perbellini F. Preparation of viable adult ventricular myocardial slices from large and small mammals. Nat Protoc. 2017;12:2623– 2639.

12. Perbellini F, Liu AKL, Watson SA, Bardi I, Rothery SM, Terracciano CM. Free-of-Acrylamide SDS-based Tissue Clearing (FASTClear) for three dimensional visualization of myocardial tissue. Sci Rep. 2017;7:5188.

13. Perbellini F, Watson SA, Scigliano M, Alayoubi S, Tkach S, Bardi I, Quaife N, Kane C, Dufton NP, Simon A, Sikkel MB, Faggian G, Randi AM, Gorelik J, Harding SE, Terracciano CM. Investigation of cardiac fibroblasts using myocardial slices. Cardiovasc Res. 2018;114:77–89.

14. Watson SA, Duff J, Bardi I, Zabielska M, Atanur SS, Jabbour RJ, Simon A, Tomas A, Smolenski RT, Harding SE, Perbellini F, Terracciano CM. Biomimetic electromechanical stimulation to maintain adult myocardial slices in vitro. Nat Commun. 2019;10:2168.

15. Nunez-Toldra R, Kirwin T, Ferraro E, Pitoulis FG, Nicastro L, Bardi I, Kit-Anan W, Gorelik J, Simon AR, Terracciano CM. Mechanosensitive molecular mechanisms of myocardial fibrosis in living myocardial slices. ESC Heart Fail. 2022;9:1400–1412.

16. Pitoulis FG, Nunez-Toldra R, Xiao K, Kit-Anan W, Mitzka S, Jabbour RJ, Harding SE, Perbellini F, Thum T, de Tombe PP, Terracciano CM. Remodelling of adult cardiac tissue subjected to physiological and pathological mechanical load *in vitro*. Cardiovasc Res. 2022;118:814–827.

17. Andreu Z, Yáñez-Mó M. Tetraspanins in Extracellular Vesicle Formation and Function. Front Immunol. 2014;5.

18. Grossman W, Paulus WJ. Myocardial stress and hypertrophy: a complex interface between biophysics and cardiac remodeling. Journal of Clinical Investigation. 2013;123:3701–3703.

19. Travers JG, Kamal FA, Robbins J, Yutzey KE, Blaxall BC. Cardiac Fibrosis. Circ Res. 2016;118:1021–1040.

20. Wencker D, Chandra M, Nguyen K, Miao W, Garantziotis S, Factor SM, Shirani J, Armstrong RC, Kitsis RN. A mechanistic role for cardiac myocyte apoptosis in heart failure. Journal of Clinical Investigation. 2003;111:1497–1504.

21. Chaitanya GV, Alexander JS, Babu PP. PARP-1 cleavage fragments: Signatures of cell-death proteases in neurodegeneration. Cell Communication and Signaling. 2010;8.

22. Mohammed SF, Hussain S, Mirzoyev SA, Edwards WD, Maleszewski JJ, Redfield MM. Coronary Microvascular Rarefaction and Myocardial Fibrosis in Heart Failure With Preserved Ejection Fraction. Circulation. 2015;131:550–559.

23. Zeng H, Chen J-X. Microvascular Rarefaction and Heart Failure With Preserved Ejection Fraction. Front Cardiovasc Med. 2019;6.

24. Huebsch N, Loskill P, Mandegar MA, Marks NC, Sheehan AS, Ma Z, Mathur A, Nguyen TN, Yoo JC, Judge LM, Spencer CI, Chukka AC, Russell CR, So P-L, Conklin BR, Healy KE. Automated Video-Based Analysis of Contractility and Calcium Flux in Human-Induced Pluripotent Stem Cell-Derived Cardiomyocytes Cultured over Different Spatial Scales. Tissue Eng Part C Methods. 2015;21:467–479.

25. Oh JG, Lee P, Gordon RE, Sahoo S, Kho C, Jeong D. Analysis of extracellular vesicle miRNA profiles in heart failure. J Cell Mol Med. 2020;24:7214–7227.

26. Loyer X, Zlatanova I, Devue C, Yin M, Howangyin K-Y, Klaihmon P, Guerin CL, Kheloufi M, Vilar J, Zannis K, Fleischmann BK, Hwang DW, Park J, Lee H, Menasché P, Silvestre J-S, Boulanger CM. Intra-Cardiac Release of Extracellular Vesicles Shapes Inflammation Following Myocardial Infarction. Circ Res. 2018;123:100–106.

27. Sahoo S, Losordo DW. Exosomes and Cardiac Repair After Myocardial Infarction. Circ Res. 2014;114:333–344.

28. Tanai E, Frantz S. Pathophysiology of Heart Failure. In: Comprehensive Physiology. Wiley; 2015. p. 187–214.

29. Pitoulis F, Watson SA, Dries E, Bardi I, Nunez-Toldra R, Perbellini F, Terracciano CM. Myocardial Slices - A Novel Platform for In Vitro Biomechanical Studies. Biophys J. 2019;116:30a.

30. Schwinger RH, Böhm M, Koch A, Schmidt U, Morano I, Eissner HJ, Uberfuhr P, Reichart B, Erdmann E. The failing human heart is unable to use the Frank-Starling mechanism. Circ Res. 1994;74:959–969.

31. Stienen GJM. Pathomechanisms in heart failure: the contractile connection. J Muscle Res Cell Motil. 2015;36:47–60.

32. Nunez-Toldra R, Kirwin T, Ferraro E, Pitoulis FG, Nicastro L, Bardi I, Kit-Anan W, Gorelik J, Simon AR, Terracciano CM. Mechanosensitive molecular mechanisms of myocardial fibrosis in living myocardial slices. ESC Heart Fail. 2022;9:1400–1412.

33. Hutcheson JD, Aikawa E. Extracellular vesicles in cardiovascular homeostasis and disease. Curr Opin Cardiol. 2018;33:290–297.

34. Gupta S, Knowlton AA. HSP60 trafficking in adult cardiac myocytes: role of the exosomal pathway. American Journal of Physiology-Heart and Circulatory Physiology. 2007;292:H3052– H3056.

35. Tian C, Gao L, Zimmerman MC, Zucker IH. Myocardial infarction-induced microRNA-enriched exosomes contribute to cardiac Nrf2 dysregulation in chronic heart failure. American Journal of Physiology-Heart and Circulatory Physiology. 2018;314:H928–H939.

36. Wang Y, Xie Y, Zhang A, Wang M, Fang Z, Zhang J. Exosomes: An emerging factor in atherosclerosis. Biomedicine & Pharmacotherapy. 2019;115:108951.

37. Li N, Rochette L, Wu Y, Rosenblatt-Velin N. New Insights into the Role of Exosomes in the Heart After Myocardial Infarction. J Cardiovasc Transl Res. 2019;12:18–27.

38. Emanueli C, Shearn AIU, Angelini GD, Sahoo S. Exosomes and exosomal miRNAs in cardiovascular protection and repair. Vascul Pharmacol. 2015;71:24–30.

39. Sahoo S, Adamiak M, Mathiyalagan P, Kenneweg F, Kafert-Kasting S, Thum T. Therapeutic and Diagnostic Translation of Extracellular Vesicles in Cardiovascular Diseases. Circulation. 2021;143:1426–1449.

40. Ganesan J, Ramanujam D, Sassi Y, Ahles A, Jentzsch C, Werfel S, Leierseder S, Loyer X, Giacca M, Zentilin L, Thum T, Laggerbauer B, Engelhardt S. MiR-378 Controls Cardiac Hypertrophy by Combined Repression of Mitogen-Activated Protein Kinase Pathway Factors. Circulation. 2013;127:2097–2106.

41. Oikawa S, Wada S, Lee M, Maeda S, Akimoto T. Role of endothelial microRNA-23 clusters in angiogenesis in vivo. American Journal of Physiology-Heart and Circulatory Physiology. 2018;315:H838–H846.

42. Xing Y, Hou J, Guo T, Zheng S, Zhou C, Huang H, Chen Y, Sun K, Zhong T, Wang J, Li H, Wang T. microRNA-378 promotes mesenchymal stem cell survival and vascularization under hypoxic– ischemic conditions in vitro. Stem Cell Res Ther. 2014;5:130.

43. Tan J, Shen J, Zhu H, Gong Y, Zhu H, Li J, Lin S, Wu G, Sun T. miR-378a-3p inhibits ischemia/reperfusion-induced apoptosis in H9C2 cardiomyocytes by targeting TRIM55 via the DUSP1-JNK1/2 signaling pathway. Aging. 2020;12:8939–8952.

44. Wu F, Wang F, Yang Q, Zhang Y, Cai K, Liu L, Li S, Zheng Y, Zhang J, Gui Y, Wang Y, Wang X, Gui Y, Li Q. Upregulation of miRNA-23a-3p rescues high glucose-induced cell apoptosis and proliferation inhibition in cardiomyocytes. In Vitro Cell Dev Biol Anim. 2020;56:866–877.

45. O’Brien J, Hayder H, Zayed Y, Peng C. Overview of microRNA biogenesis, mechanisms of actions, and circulation. Front Endocrinol (Lausanne). 2018;9.

46. Kehat I, Molkentin JD. Molecular Pathways Underlying Cardiac Remodeling During Pathophysiological Stimulation. Circulation. 2010;122:2727–2735.

47. Pitoulis FG, Terracciano CM. Heart Plasticity in Response to Pressure- and Volume-Overload: A Review of Findings in Compensated and Decompensated Phenotypes. Front Physiol. 2020;11.

48. Gobeil S, Boucher CC, Nadeau D, Poirier GG. Characterization of the necrotic cleavage of poly(ADP-ribose) polymerase (PARP-1): implication of lysosomal proteases. Cell Death Differ. 2001;8:588–594.

49. Luxán G, Dimmeler S. The vasculature: a therapeutic target in heart failure? Cardiovasc Res. 2022;118:53–64.

50. Brutsaert DL. Cardiac Endothelial-Myocardial Signaling: Its Role in Cardiac Growth, Contractile Performance, and Rhythmicity. Physiol Rev. 2003;83:59–115.

51. Ahmed RE, Anzai T, Chanthra N, Uosaki H. A Brief Review of Current Maturation Methods for Human Induced Pluripotent Stem Cells-Derived Cardiomyocytes. Front Cell Dev Biol. 2020;8.

52. Hua Z, Lv Q, Ye W, Wong CKA, Cai G, Gu D, Ji Y, Zhao C, Wang J, Yang BB, Zhang Y. Mirna-directed regulation of VEGF and other angiogenic under hypoxia. PLoS One. 2006;1.

53. Krist B, Podkalicka P, Mucha O, Mendel M, Sępioł A, Rusiecka OM, Józefczuk E, Bukowska-Strakova K, Grochot-Przęczek A, Tomczyk M, Klóska D, Giacca M, Maga P, Niżankowski R, Józkowicz A, Łoboda A, Dulak J, Florczyk-Soluch U. miR-378a influences vascularization in skeletal muscles. Cardiovasc Res. 2020;116:1386–1397.

